# Bidirectional association and shared risk factors between atrial fibrillation and heart failure: a Mendelian Randomization study

**DOI:** 10.1101/2023.08.21.23294384

**Authors:** Haibin Lu, Ziting Gao, Hongye Wei, Yajing Wei, Ziyi Qiu, Jun Xiao, Wuqing Huang

## Abstract

**Background:** Atrial fibrillation and heart failure are closely related and share multiple risk factors. We aimed to apply the mendelian randomization (MR) analysis to explore the bidirectional causal link between atrial fibrillation and heart failure, and the independent effect of potential risk factors on the risk of both conditions.

**Methods:** This is a two-sample MR study using publicly available summary-level statistics of genome-wide association studies (GWAS). Bidirectional MR was performed to explore the relation between atrial fibrillation and heart failure. A total of 14 factors were selected as potential risk factors, univariable MR analyses were used to identify shared risk factors, and then the multivariable MR analyses were further used to investigate the independent effect of these factors on both conditions. Inverse-variance-weighted MR (IVW-MR) were used to obtain the effect estimates.

**Results:** MR analysis found evidence of causal relationship between atrial fibrillation and heart failure (odds ratio [OR], 1.24; 95% confidence interval [CI], 1.19–1.29), as well as between heart failure and atrial fibrillation (OR, 3.88; 95% CI, 1.45–10.37). Univariable MR analyses identified several shared risk factors for both conditions, including body mass index (BMI), blood pressure, smoking, coronary heart disease and myocardial infarction. After adjusting for atrial fibrillation, the observed associations between shared factors and heart failure kept stable, such as BMI, smoking, coronary heart disease and myocardial infarction. However, after adjusting for heart failure, the relationships between most risk factors and atrial fibrillation attenuated to null.

**Conclusions:** This two-sample MR study found a bidirectional relationship between atrial fibrillation and heart failure, and identified several shared risk factors of both conditions, which had an independent effect on the risk of heart failure while probably affected the risk of atrial fibrillation via cardiac impairment.

**Funding:** Start-up Fund for high-level talents of Fujian Medical University (grant no.XRCZX2021026) and Natural Science Foundation of Fujian Province (grant no. 2022J01706).

## Introduction

Atrial fibrillation and heart failure have emerged as a dual epidemic, together resulting in substantial public health burden (**Anter E et al., 2009**). Atrial fibrillation is the most common clinical arrhythmia with an estimation of 4.7 million incident cases and 60 million prevalent cases globally in 2019. And the number of atrial fibrillation cases is expected to double by 2060, showing that atrial fibrillation remains a major public health concern (**Dong XJ et al., 2023**). And the prevalence of heart failure in adults was reported to increase from 1-2 percent in adults to more than 10 percent in elderly population over 70 years of age (**Jia Q et al., 2019**). With the increasing life-expectancy of the general population, heart failure becomes currently the fastest-growing cardiovascular disease worldwide with a rough estimation of more than 64.3 million cases globally in 2017, which has been defined as a global pandemic (**Savarese G et al., 2023**).

The coexistence of atrial fibrillation and heart failure has been appreciated for several decades (**Turagam MK et al., 2019**). Previous studies have shown that persistent or frequent atrial fibrillation would lead to irreversible cardiac structural changes, resulting in impairment of systolic and diastolic function, and further increasing the incidence of heart failure (**Carlisle MA et al., 2019**). Most patients with heart failure suffered atrial enlargement, mitral regurgitation, and changes in neurohumoral balance, all of which are related to higher risk of atrial fibrillation (**Verhaert DVM et al., 2021**). However, it is unclear regarding the causative relationship between atrial fibrillation and heart failure. In addition, as reported in previous observational studies, atrial fibrillation and heart failure share a range of risk factors, including hypertension, diabetes, structural heart disease, smoking, alcoholic drinking, obesity and so on (**Kornej J et al., 2021; Young LJ et al., 2022; Meijers WC et al., 2019; Roger VL, 2021**). While lack of studies distinguished the independent effect of these common risk factors on the risk of atrial fibrillation or heart failure, and mediated role of one condition between common risk factor and the other condition.

Therefore, in this study, we aimed to apply the mendelian randomization (MR) analysis to explore the bidirectional causal link between atrial fibrillation and heart failure, and the independent effect of potential risk factors on the risk of both conditions. MR study use genetic variation closely related to the exposure as instrumental variables to infer causal associations between exposure and outcome events (**Zheng J et al., 2017**). Using genetic variation as a tool can effectively reduce the impact of confounding factors and avoid reverse causality. In this study, bidirectional MR analysis was first conducted to explore the bidirectional association between atrial fibrillation and heart failure; next, univariable MR analysis was performed to identify the potential shared risk factors of both conditions, and multivariable MR analysis including each shared risk factor and both conditions was further applied to explore the effect of shared risk factors on the risk of one condition independent of the other one, as well as assess the mediated role of one condition between the factor of interest and risk of the other one.

## Methods

### Data sources

Publicly available summary statistics for genetic variants related to exposures and outcomes were obtained from large-scaled genome-wide association studies (GWAS) of European participants and used to perform two-sample MR study (**Supplementary File-Table 1**). All these studies had been approved by the relevant institutional review boards and informed consent had been obtained for all participants per the original study protocols.

Summary-level GWAS data for atrial fibrillation were derived from a meta-analysis consisting of six large cohorts: The Nord-Trøndelag Health Study, deCODE, the Michigan Genomics Initiative, DiscovEHR Collaboration Cohort, UK Biobank and AFGen Consortium. This study included 60,620 individuals with atrial fibrillation and 970,216 controls (**Nielsen JB et al., 2018**). Cases of atrial fibrillation were mainly identified by ICD-9 (427.3) or ICD-10 (I48) codes (**Nielsen JB et al., 2018**). Summary-level GWAS data for heart failure were obtained from the Heart Failure Molecular Epidemiology for Therapeutic Targets (HERMES) Consortium consisting of 26 European population cohorts, including 47,309 cases of heart failure and 930,014 controls (**Shah S et al., 2018**).

Based on the previous literature, this study summarized several potential common risk factors for atrial fibrillation and heart failure, and classified these risk factors into three categories: metabolic trait (waist circumference, body mass index [BMI], systolic blood pressure, diastolic blood pressure, glycosylated hemoglobin type A1C [HbA1C], fasting glucose, fasting insulin), environmental or behavior factor (smoking, alcoholic drinking, physical activity, particulate matter 2.5 [PM2.5] exposure) and comorbidity (coronary heart disease, myocardial infarction). Summary statistics were obtained from GIANT consortium for waist circumference (n=245,746) and body mass index (n=681,275), from ICSB consortium for blood pressure (n=757,601), from MAGIC consortium for HbA1C (n=46,368), fasting glucose (n=58,074) and fasting insulin (n=51,750), from GSCAN consortium for smoking (ncases=311,629; ncontrols=321,173) and alcoholic drinking (n=335,394), from UK Biobank for physical activity (n=440,512) and PM2.5 exposure (n=423,796), from CARDIoGRAMplusC4D consortium for coronary heart disease (ncases=60,801; ncontrols=123,504) and myocardial infarction (ncases=43,676; ncontrols=128,199). Detailed information of data sources used in the analysis are provided in **Supplementary File-Table 1.**

### Selection of genetic instrumental variables

Single nucleotide polymorphisms (SNPs) which met genomic significance threshold (p<5*10^-8^) were selected from the GWAS pooled database as genetic instrument**s** corresponding to the exposure of interest (**Sanna S et al., 2019**). Linkage disequilibrium was estimated by R^2^<0.001 (clumping window size=10,000kb) using genomic data from the European population in the 1000 Genomes Project as the reference dataset, and SNP with strongest association with the exposure was retained if linkage disequilibrium was present. For SNPs unavailable in the GWAS dataset of outcome, suitable proxies were used as substitutes, SNPs without suitable proxies and palindromic SNPs were removed. Then remaining independent SNPs with statistically significance were used as genetic instruments.

### Statistical analysis

#### Primary analyses

As shown in flowchart in **Fig 1**, bidirectional MR analysis was first performed to explore the causal association between atrial fibrillation and heart failure. Next, we performed a univariable MR analysis to estimate the causal effect of potential factors on the risk of atrial fibrillation and heart failure respectively, then identified the shared risk factors between atrial fibrillation and heart failure. Finally, multivariable MR analysis was conducted to investigate the impact of each shared risk factor on one outcome independent of the other outcome. The random-effect inverse-variance weighted mendelian randomization (IVW-MR) was used as the main method (**Burgess S et al., 2013**). In MR analysis, we used the F-statistic to assess the strength of exposed genetic tools, and F-statistic >10 was not considered to be biased by weak genetic tools (**Sanderson E et al., 2015; Burgess S et al., 2011**).

**Fig 1:**
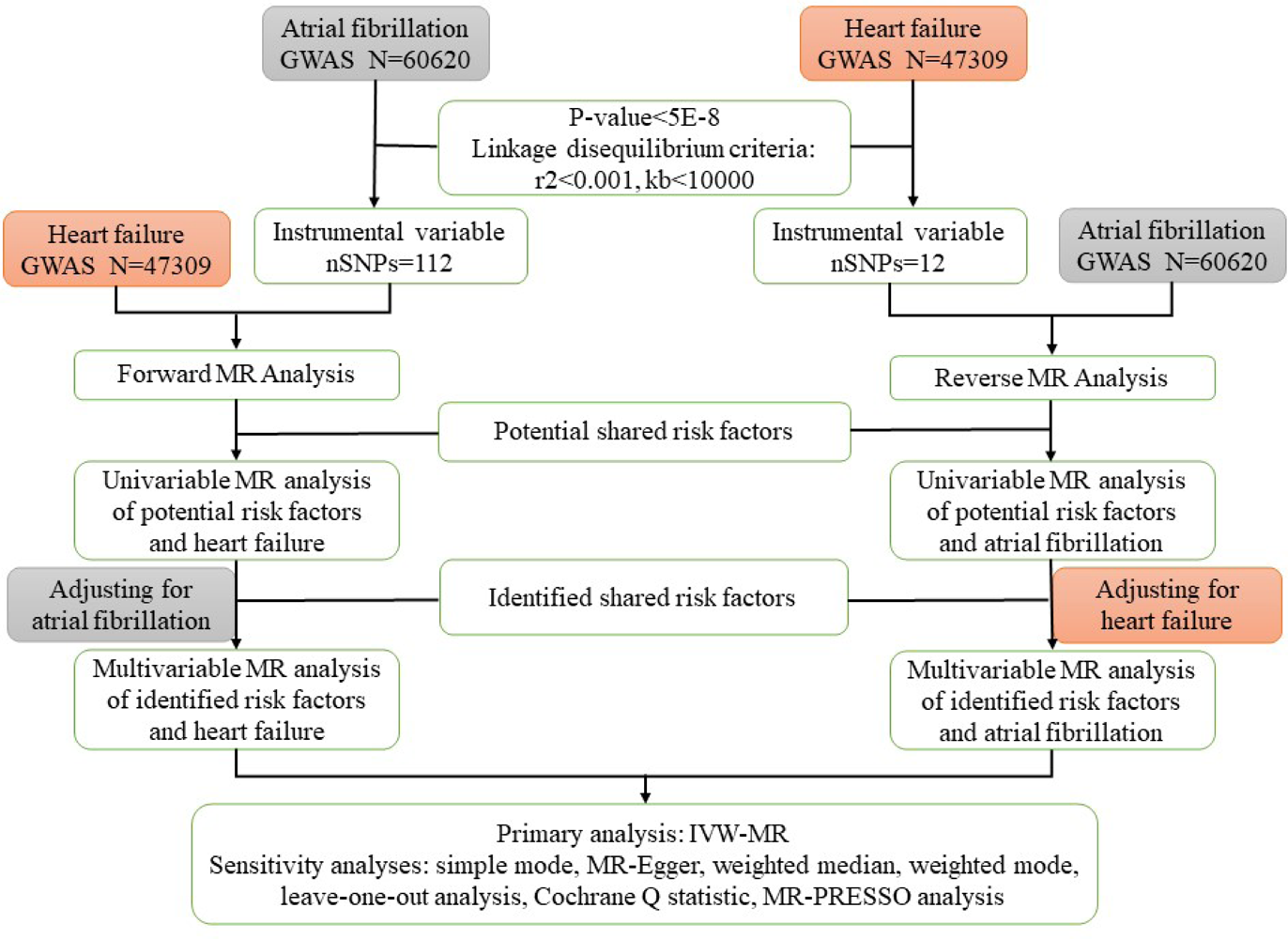
A flow chart of the study design.

To account for multiple testing, Bonferroni correction was used to adjust the thresholds of significance level, thus a strong evidence was suggested for p<0.002 (thirteen exposures and two outcomes) and a suggestive evidence of 0.002≤ p< 0.05 in univariable MR. And a strong evidence was suggested for p<0.004 (six exposures and two outcomes) and a suggestive evidence of 0.004≤ p<0.05 in multivariable MR.

#### Sensitivity analyses

Several MR methods were used to examine the robustness of the main results, including simple mode, MR-Egger, weighted median, weighted mode. Sensitivity analyses, including the leave-one-out analysis, Cochran’s Q statistic, MR-Egger intercept and MR-pleiotropy residual sum and outlier (MR-PRESSO), were applied to assess the possible heterogeneity and horizontal pleiotropy in the primary analyses. The leave-one-out analysis refers to evaluate the consistency of observed associations by removing one SNP from analysis in turn and identify whether the results were driven by any individual SNP. The Cochrane Q statistic was used for heterogeneity in this study, and a Cochran’s Q-derived P<0.05 was deemed as a marker of the presence of heterogeneity. The MR-Egger intercept and MR-PRESSO analysis were used to evaluate the horizontal pleiotropy of the instrumental variables. The MR-PRESSO analysis consists of three components (**Verbanck M et al., 2018**): (1) Detection of horizontal pleiotropy; (2) Correction of pleiotropy by removing detected outliers (genetic variants with horizontal pleiotropy); (3) Comparison of the differences in causal correlation before and after correction.

All statistical analyses were performed using R version 4.2.2 (R Foundation for Statistical Computing, Vienna, Austria). MR analyses were performed using the TwosampleMR, MRPRESSO and MRInstruments R packages.

## Results

### Bidirectional MR analysis of atrial fibrillation and heart failure

We used a two-way two-sample MR study to explore the bidirectional relationship between atrial fibrillation and heart failure. The F-statistic for atrial fibrillation and heart failure were all greater than 10 (**Supplementary File-Table 1 and Table 2**). We found evidence of causal relationship between atrial fibrillation and heart failure (odds ratio [OR], 1.24; 95% confidence interval [CI], 1.19–1.29; P<0.001), as well as between heart failure and atrial fibrillation (OR, 3.88; 95% CI, 1.45–10.37; P=0.007) (**Fig 2**). Four complementary methods yielded similar results between atrial fibrillation and heart failure, and consistent evidence for the causal relationship of heart failure with atrial fibrillation was found across these methods (**Fig 2**). The scatterplot provides an estimate of one condition for each genetic variant to the other one (**Supplementary File-Fig 1**). The leave-one-out analysis of the MR association did not find any SNP that had a significant impact on the estimated effect (**Supplementary File-Fig 2**). The Cochrane Q statistic showed an evidence of heterogeneity (P<0.001), thus random-effect IVW-MR was used as the main method (**Supplementary File-Table 4**). Potential horizontal pleiotropy was not found in the genetic instruments of atrial fibrillation (MR-Egger intercept P =0.171; MR-PRESSO P=1.000) and heart failure (MR-Egger intercept P =0.090; MR-PRESSO P=0.675; **Supplementary File-Table 4**).

**Fig 2:**
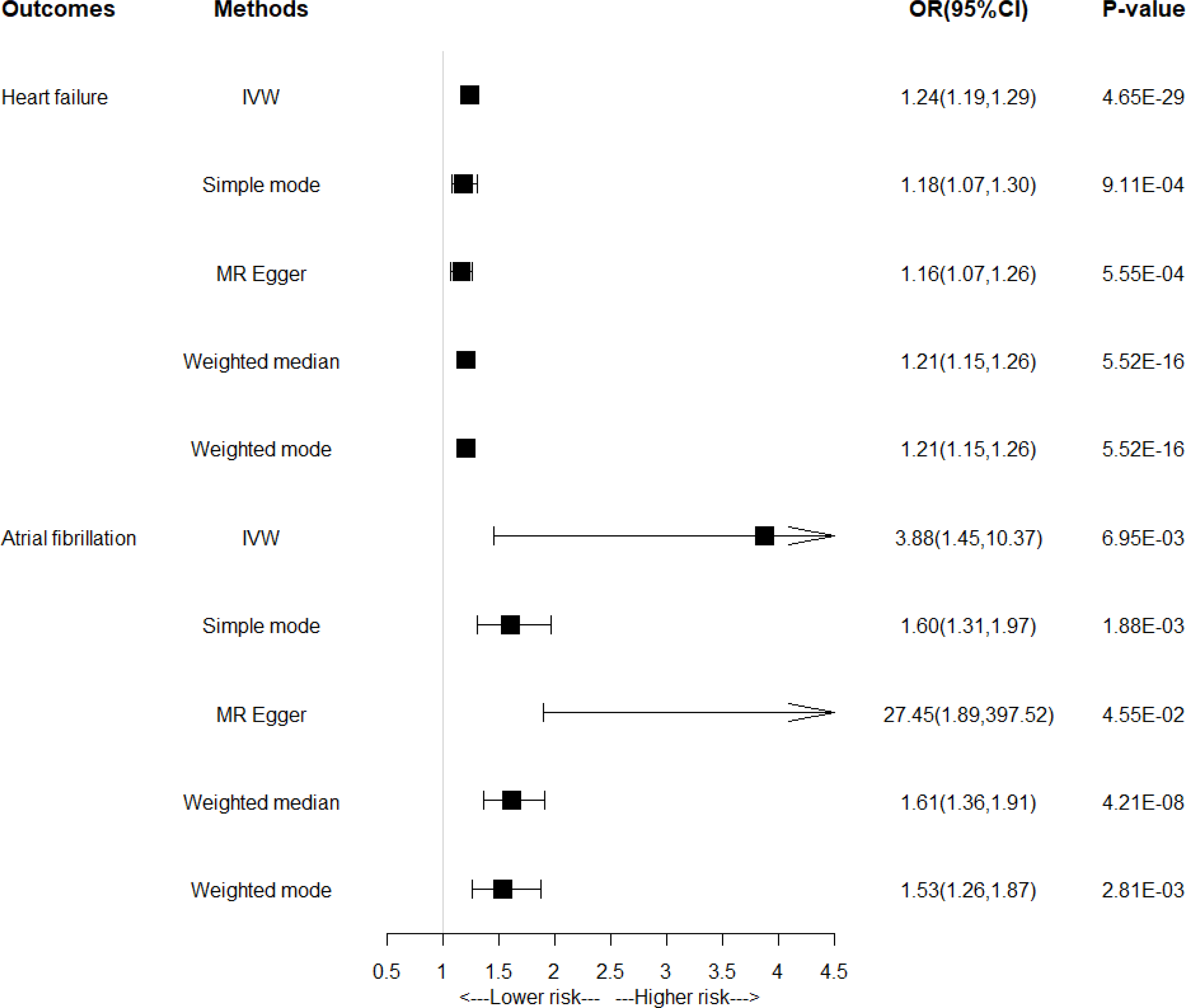
Bidirectional MR analysis between atrial fibrillation and heart failure.

### Univariable MR analysis for risk factors on atrial fibrillation and heart failure

As shown in **Fig 3**, a strong evidence was found between several factors and both conditions, including BMI (heart failure: OR, 1.74; 95% CI, 1.63–1.85; P<0.001; atrial fibrillation: OR, 1.41; 95% CI, 1.33–1.50; P<0.001), systolic blood pressure (heart failure: OR, 1.02; 95% CI, 1.02–1.03; P<0.001; atrial fibrillation: OR, 1.02; 95% CI, 1.01–1.02; P<0.001), diastolic blood pressure (heart failure: OR, 1.03; 95% CI, 1.02–1.04; P<0.001; atrial fibrillation: OR, 1.03; 95% CI, 1.02–1.04; P<0.001), smoking (heart failure: OR, 1.27; 95% CI, 1.15–1.39; P<0.001; atrial fibrillation: OR, 1.09; 95% CI, 1.01–1.18; P=0.0308), coronary heart disease (heart failure: OR, 1.34; 95% CI, 1.27–1.41; P<0.001; atrial fibrillation: OR, 1.11; 95% CI, 1.05–1.17; P<0.001) and myocardial infarction (heart failure: OR, 1.35; 95% CI, 1.26–1.44; P<0.001; atrial fibrillation: OR, 1.11; 95% CI, 1.06–1.16; P<0.001). Heterogeneity may be present in the instrumental variables for most exposures, thus random-effect IVW-MR was used as the main method (**Supplementary File-Table 4**). And the MR-Egger intercept and the MR-PRESSO analysis showed the presence of potential horizontal pleiotropy in the instrumental variables for myocardial infarction (MR-Egger intercept P=0.039), while there was no horizontal pleiotropy in other exposure factors (**Supplementary File-Table 4**).

**Fig 3.**
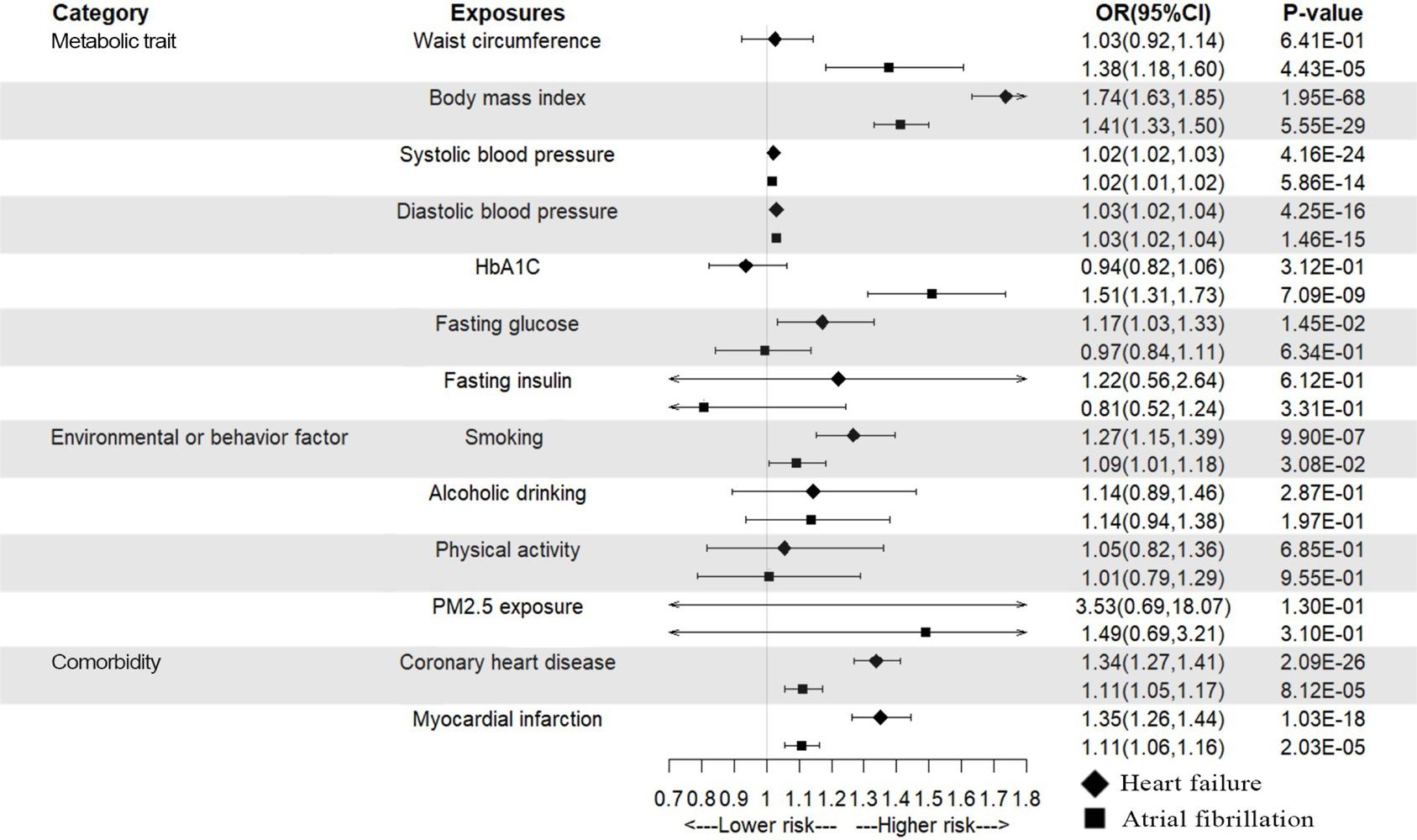
Univariable MR analysis of potential shared risk factors with atrial fibrillation and heart failure.

Univariable MR analysis also found a suggestive evidence for the association of fasting glucose with the higher risk of heart failure (OR, 1.17; 95% CI, 1.03–1.33; P=0.0145); a strong evidence for waist circumference (OR, 1.38; 95% CI, 1.18–1.60; P<0.001) and HbA1C (OR, 1.51; 95% CI, 1.31–1.73; P<0.001) were correlated with the higher risk of atrial fibrillation; and no evidence for between fasting insulin, alcoholic drinking, physical activity and PM2.5 exposure were correlated with both heart failure and atrial fibrillation (**Fig 3**). The Cochrane Q statistic showed an evidence of heterogeneity among the instrument SNP effects except for fasting glucose (P=0.718) and fasting insulin (P=0.316; **Supplementary File-Table 4**). And the MR-PRESSO analysis showed the presence of potential horizontal pleiotropy in the instrumental variables of waist circumference (P=0.008), physical activity (P=0.029) and PM2.5 exposure (P<0.001; **Supplementary File-Table 4**).

### Multivariable MR analysis for shared risk factors on atrial fibrillation and heart failure

To obtain the independent effect of shared risk factors on each condition, we used the multivariable MR approach to adjust for the effect from the other condition. As shown in **Fig 4**, after adjusting for atrial fibrillation, the observed associations between shared factors and heart failure kept stable, such as BMI (OR, 1.58; 95% CI, 1.49– 1.68), smoking (OR, 1.24; 95% CI, 1.10–1.41), coronary heart disease (OR, 1.29; 95% CI, 1.23–1.36) and myocardial infarction (OR, 1.30; 95% CI, 1.22–1.38). However, after adjusting for heart failure, the relationships between most risk factors and atrial fibrillation attenuated to null, even a negative association was found between coronary heart disease (OR, 0.72; 95% CI, 0.59–0.88) or myocardial infarction (OR, 0.69; 95% CI, 0.55–0.88) and atrial fibrillation.

**Fig 4.**
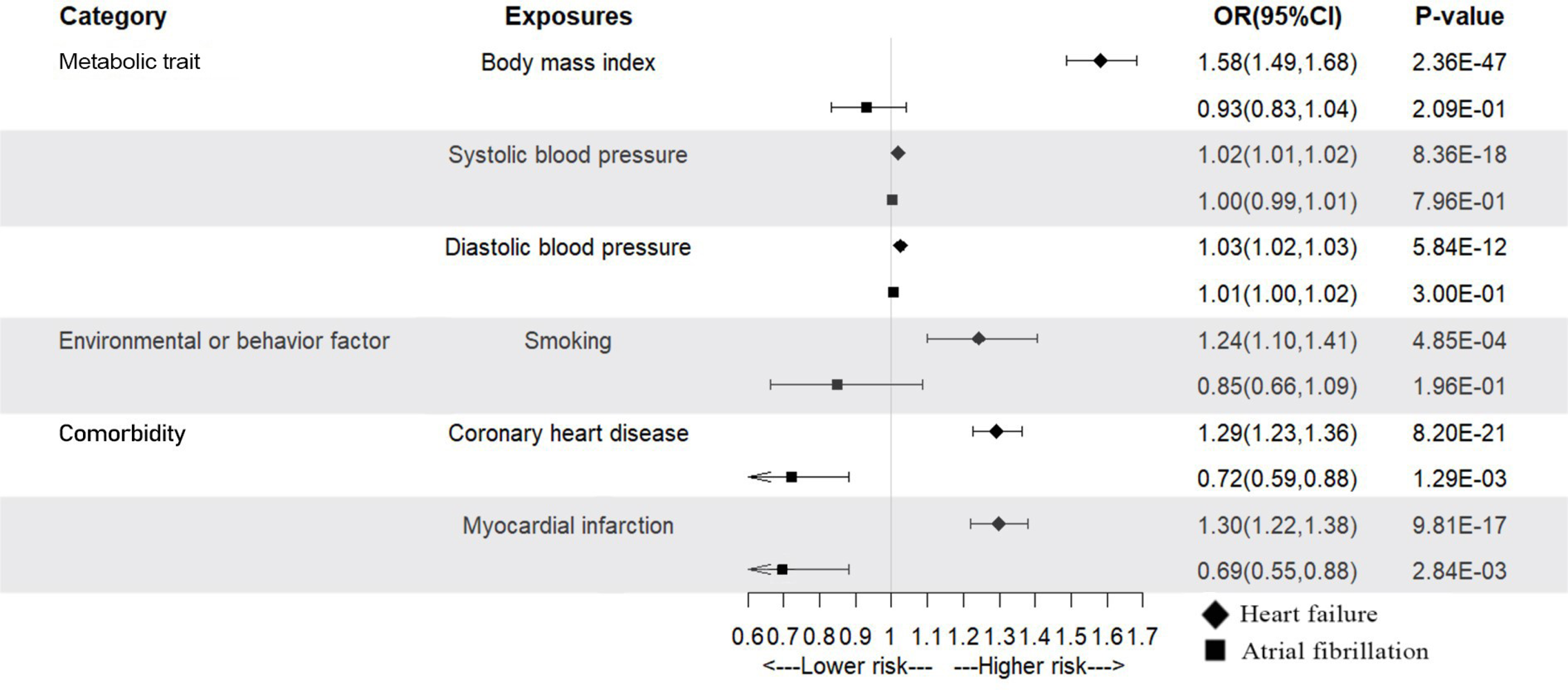
Multivariable MR analysis of identified shared risk factors with atrial fibrillation and heart failure.

## Discussion

In the present study, a significant bidirectional association was found between atrial fibrillation and heart failure, in which atrial fibrillation were related to 1.24 times higher risk of heart failure, and heart failure associated with 3.88 times higher risk of atrial fibrillation. BMI, blood pressure, smoking, coronary heart disease and myocardial infarction were identified as shared risk factors of atrial fibrillation and heart failure. The observed associations between shared risk factors and heart failure remained stable after adjusting for atrial fibrillation, suggesting that these shared risk factors were independently related to heart failure risk. However, after adjusting for heart failure, the observed associations between most shared risk factors and atrial fibrillation attenuated to null, indicated that heart failure played an important mediated role between shared risk factors and risk of atrial fibrillation. These findings suggested that weight control, blood pressure control and smoking quitting might reduce the risk of heart failure, and individuals with cardiac impairment were likely to benefit from these strategies in reducing atrial fibrillation risk.

A number of epidemiological studies suggested a link between atrial fibrillation and heart failure (**Anter E et al., 2009**), as reported, the incidence of atrial fibrillation in heart failure patients was as high as 20% to 60% (**Ponikowski P et al., 2021; Muser D et al., 2019; Zakeri R et al., 2013; Prabhu S et al., 2017**), and the risk of developing heart failure in atrial fibrillation patients increased by five to six times (**Ruddox V et al., 2017; Vermond RA et al., 2015**). However, due to the chronic nature of both conditions, it is difficult to avoid the impact of reverse causality in the observational studies when estimating the causative relationship between atrial fibrillation and heart failure. In recent years, some MR studies have explored the role of atrial fibrillation on the risk of heart failure (**Shah S et al., 2020; Kwok MK et al., 2021; Hu M et al., 2023**), but there is a dearth of evidence from MR study fully exploring the bidirectional causal relationship between these two conditions. The present bidirectional MR study provided additional evidence for the link between both conditions, showing that atrial fibrillation was related to 1.24 times higher risk of heart failure, and heart failure associated with 3.88 times higher risk of atrial fibrillation, further confirming the important influence of atrial fibrillation and heart failure on each other’s occurrence and development.

A series of common risk factors have been related to both atrial fibrillation and heart failure (**Kornej J et al., 2021; Young LJ et al., 2022; Meijers WC et al., 2019; Roger VL, 2021**), the coexistence of both conditions may be explained by the presence of these common risk factors to some extent, but lack of studies comprehensively investigated how these risk factors independently affect each condition, respectively. In this study, univariable MR analyses identified several shared risk factors for both diseases, including BMI, blood pressure, smoking, coronary heart disease and myocardial infarction. The results are consistent with those of Susanna et al. who found a causal relationship between BMI and smoking with atrial fibrillation and heart failure (**Larsson SC et al., 2020; Larsson SC et al., 2020**), and Nhu et al. who found a causal relationship between blood pressure with atrial fibrillation and heart failure (**Le NN et al., 2022**). Due to the bidirectional relationship between atrial fibrillation and heart failure, we further performed multivariable MR analyses to investigate the effect of these shared factors on each disease independent on the other one in this study, in which we have found a strong evidence that BMI, blood pressure, smoking, coronary heart disease and myocardial infarction were independent risk factors for heart failure, but not for atrial fibrillation. After adjusting for heart failure, the relationship of most shared factors (eg, BMI, blood pressure, smoking) with atrial fibrillation was basically not significant, indicating that these shared risk factors probably increased the risk of atrial fibrillation via impairment in cardiac structure and function. These findings suggested that individuals with cardiac impairment were likely to benefit from weight control, blood pressure control and smoking quitting in reducing atrial fibrillation risk. Interestingly, after adjusting for heart failure, this study found a negative correlation between coronary heart disease or myocardial infarction and atrial fibrillation. Similarly, Bouwe et al. previously reported a negative correlation between myocardial infarction and atrial fibrillation in women (hazard ratio [HR], 0.92; 95%CI, 0.59–1.44) (**Krijthe BP et al., 2013**), the underlying reason of which remained to be clarified.

This study has several advantages. To our knowledge, this is the first study to explore the bidirectional relationship between atrial fibrillation and heart failure from a genetic perspective, identify shared risk factors of both conditions and the independent effect of shared risk factors on each outcome. The bidirectional causal relationship between atrial fibrillation and heart failure was determined through MR analysis, minimizing confounding impact and reverse causal relationships. In addition, this study is the first to simultaneously explore the possible effect of metabolic trait, environmental or behavior factor and comorbidity in the occurrence of atrial fibrillation and heart failure at the genetic level, comprehensively exploring whether there are differences in the strength of the impact of different risk factors on atrial fibrillation and heart failure. To ensure the reliability of the results, we have adopted a series of supplementary methods and sensitivity analyses.

However, this study also has some limitations. Firstly, in the GWAS study of atrial fibrillation and heart failure, the aggregated data come from multiple different cohorts, and their definitions of atrial fibrillation and heart failure vary, which to some extent weakens the causal effects of related genetic tools. However, the studies provide a comparison of the direction of genetic variation effects significantly associated with previous heart failure and atrial fibrillation, and the large sample size also ensures consistency in the direction of genetic effects (**Shah S et al., 2020; Nielsen JB et al., 2018**). Secondly, the GWAS summary data used in this study were obtained from relevant studies based on European populations, so the generalization of the research results is limited and further testing is needed in other ethnic groups. Third, potential pleiotropy cannot be ruled out, which might have biased the results. Nonetheless, several approaches were performed to minimize the probability of pleiotropy bias, including identifying pleiotropy with MR-Egger intercept and detecting outliers by the MR-PRESSO analysis. Fourthly, the MR analysis of this study is based on aggregated data and cannot further explore whether there are differences in associations among different subgroups of the population at the individual level.

## Conclusions

In summary, this two-sample MR study found a bidirectional relationship between atrial fibrillation and heart failure, and identified several shared risk factors of both conditions, including BMI, blood pressure, smoking, coronary heart disease and myocardial infarction, all of which were independently related to heart failure but affected the risk of atrial fibrillation probably via impairment in cardiac structure and function.

## Acknowledgments

We thank the patients and investigators who contributed to the UK Biobank, GIANT Consortium, HERMES Consortium, ICBP consortium, MAGIC consortium, GSCAN Consortium, CARDIoGRAMplusC4D Consortium.

## Author Contributions

All authors were responsible for the study concept and design. WH obtained funding. HL and ZG did the statistical analysis and drafted the manuscript. WH and JX critically revised the manuscript. All authors gave the final approval and agreed to be accountable for all aspects of work ensuring integrity and accuracy.

## Data Availability Statement

All raw summary-level data are available in the referenced public datasets, the detailed information of which could be found in Supplementary File-Table 1. The codes and curated data for the current analysis are available at https://github.com/WH57/Atrial-fibrillation-and-heart-failure.

## Funding Information

This work was supported by the Start-up Fund for high-level talents of Fujian Medical University (grant no.XRCZX2021026) and the Natural Science Foundation of Fujian Province (grant no. 2022J01706) to Dr. Wuqing Huang. The funder had no role in study design, data collection and interpretation, or the decision to submit the work for publication.

## Declaration of Competing Interest

The authors declare no competing interests.

## Abbreviations

BMI: body mass index
CI: confidence interval
GWAS: Genome-Wide Association Studies
HbA1C: Glycosylated hemoglobin type A1C
IVW-MR: inverse-variance weighted mendelian randomization
MR: Mendelian randomization
MR-PRESSO: MR-pleiotropy residual sum and outlier
OR: odds ratio
PM2.5: Particulate matter 2.5
SNPs: single nucleotide polymorphisms.

## Supporting information

**Supplementary File.** Contents of Table 1-4 and Fig 1-2.

### Table of contents

**Supplementary File-Table 1.** Information of data sources used in the MR study.

| Traits | Data sources | Sample size<br>(cases/controls) | Number<br>of SNPs | Ancestry | Reference |
| --- | --- | --- | --- | --- | --- |
| Atrial fibrillation | Meta analysis of six cohort studies | 60,620/970,216 | 33,519,037 | European | [1] |
| Heart failure | HERMES consortium | 47,309/930,014 | 7,773,021 | European | [2] |
| Waist circumference | GIANT consortium | 245,746 | 2,547,573 | Mixed (76% European) | [3] |
| Body mass index | GIANT consortium | 681,275 | 2,336,260 | Mixed (76% European) | [4] |
| Systolic blood pressure | ICBP consortium | 757,601 | 7,088,083 | European | [5] |
| Diastolic blood pressure | ICBP consortium | 757,601 | 7,160,619 | European | [5] |
| HbA1C | MAGIC consortium | 46,368 | 2,529,804 | European | [6] |
| Fasting glucose | MAGIC consortium | 58,074 | 2,625,495 | European | [7] |
| Fasting insulin | MAGIC consortium | 51,750 | 2,624,472 | European | [7] |
| Smoking | GSCAN consortium | 311,629/321,173 | 11,802,365 | European | [8] |
| Alcoholic drinking | GSCAN consortium | 335,394 | 11,887,865 | European | [8] |
| Physical activity | UK Biobank | 440,512 | 9,851,867 | European | [9] |
| PM2.5 exposure | UK Biobank | 423,796 | 9,851,867 | European | [10] |
| Coronary heart disease | CARDIoGRAMplusC4D consortium | 60,801/123,504 | 9,455,779 | Mixed (77% European) | [11] |
| Myocardial infarction | CARDIoGRAMplusC4D consortium | 43,676/128,199 | 9,289,492 | Mixed (77% European) | [11] |

---

**Supplementary File-Table 2.** SNPs used to construct the instruments for MR analysis atrial fibrillation and heart failure (nSNPs=112).

| SNP | Chromosome | Position | Effect allele | Other allele | Exposure: Atrial fibrillation |  |  | Outcome: Heart failure |  |  | F-statistic |
| --- | --- | --- | --- | --- | --- | --- | --- | --- | --- | --- | --- |
|  |  |  |  |  | Beta | Standard error | P value | Beta | Standard error | P value |  |
| rs284277 | 1 | 10790797 | A | C | -0.0422 | 0.0069 | 1.25E-09 | -0.0139 | 0.0082 | 0.0905191 | 868.0021 |
| rs10753933 | 1 | 203026214 | G | T | -0.0609 | 0.0067 | 9.84E-20 | -0.0106 | 0.0078 | 0.1767 | 1894.542 |
| rs2540949 | 2 | 65284231 | T | A | -0.0659 | 0.0068 | 2.95E-22 | -0.0067 | 0.008 | 0.4041 | 2123.697 |
| rs10520260 | 4 | 174447349 | G | A | -0.0457 | 0.0073 | 3.36E-10 | 0.002 | 0.0085 | 0.8136 | 939.9542 |
| rs10741807 | 11 | 20011445 | C | T | -0.0729 | 0.0079 | 1.59E-20 | 0.0048 | 0.0093 | 0.6076 | 2030.119 |
| rs28439930 | 5 | 173393111 | C | G | -0.0458 | 0.0068 | 1.19E-11 | -0.0134 | 0.0092 | 0.146 | 1080.997 |
| rs6891790 | 5 | 172670745 | T | G | -0.0729 | 0.0076 | 4.53E-22 | -0.033 | 0.0088 | 1.81E-04 | 2227.054 |
| rs11598047 | 10 | 105342672 | G | A | 0.1537 | 0.009 | 8.95E-66 | 0.0188 | 0.0107 | 0.0774105 | 6657.906 |
| rs74910854 | 7 | 74110705 | G | A | 0.09 | 0.0164 | 4.31E-08 | 0.0587 | 0.0207 | 4.59E-03 | 1078.203 |
| rs60902112 | 3 | 194800853 | T | C | 0.0445 | 0.0079 | 1.72E-08 | 0.0056 | 0.0094 | 0.5483 | 715.0909 |
| rs8088085 | 18 | 48708548 | C | A | -0.0365 | 0.0067 | 4.79E-08 | -0.0253 | 0.0078 | 0.00121 | 683.6754 |
| rs72811294 | 17 | 12618680 | C | G | -0.072 | 0.0106 | 9.67E-12 | -0.0239 | 0.0146 | 0.1015 | 1073.181 |
| rs133885 | 22 | 26159289 | A | G | 0.0405 | 0.0068 | 2.22E-09 | 0.0136 | 0.008 | 0.0872208 | 832.9601 |
| rs1458038 | 4 | 81164723 | T | C | 0.0434 | 0.0072 | 1.74E-09 | 0.0165 | 0.0085 | 0.0530994 | 829.3745 |
| rs10520002 | 5 | 127819132 | A | G | 0.0626 | 0.0113 | 2.85E-08 | 0.0325 | 0.0135 | 0.01595 | 719.8611 |
| rs6580277 | 5 | 142818123 | G | A | 0.067 | 0.0079 | 1.64E-17 | 0.0307 | 0.0093 | 9.59E-04 | 1675.793 |
| rs4965430 | 15 | 99268850 | G | C | -0.0441 | 0.0069 | 1.26E-10 | -0.0135 | 0.0081 | 0.0942106 | 951.5225 |
| rs2834618 | 21 | 36119111 | G | T | -0.0944 | 0.0112 | 3.41E-17 | -0.0236 | 0.013 | 0.0697493 | 1738.161 |
| rs73366713 | 6 | 16415751 | A | G | -0.1035 | 0.0099 | 1.53E-25 | -0.0012 | 0.0116 | 0.9145 | 2659.526 |
| rs1545300 | 1 | 112464004 | T | C | -0.0558 | 0.0073 | 1.48E-14 | -0.0135 | 0.0085 | 0.1116 | 1372.711 |
| rs146518726 | 1 | 51535039 | A | G | 0.1605 | 0.0207 | 8.27E-15 | 0.0836 | 0.0238 | 4.50E-04 | 1687.599 |
| rs35544454 | 2 | 213266003 | T | A | -0.0589 | 0.0087 | 1.10E-11 | -0.02 | 0.0101 | 0.0468997 | 1109.901 |
| rs7696743 | 4 | 114393950 | G | A | 0.041 | 0.0069 | 2.21E-09 | 0.0293 | 0.0081 | 2.89E-04 | 808.3896 |
| rs6854883 | 4 | 111607315 | T | C | 0.2822 | 0.0093 | 1.00E-200 | 0.0594 | 0.0108 | 3.67E-08 | 21355.49 |
| rs62254082 | 3 | 69417585 | C | T | 0.0404 | 0.007 | 6.34E-09 | 0.0117 | 0.0082 | 0.152 | 798.5127 |
| rs67969609 | 2 | 145760353 | G | C | 0.0711 | 0.0126 | 1.71E-08 | 0.0374 | 0.015 | 0.0128 | 687.8943 |
| rs7574892 | 2 | 175512820 | A | G | 0.0552 | 0.0067 | 1.98E-16 | 0.0057 | 0.0078 | 0.4633 | 1571.418 |
| rs117984853 | 6 | 149399100 | T | G | 0.1228 | 0.012 | 1.34E-24 | 0.0027 | 0.0137 | 0.8437 | 2838.139 |
| rs7612445 | 3 | 179172979 | T | G | 0.0493 | 0.0084 | 4.81E-09 | 0.0047 | 0.0101 | 0.642899 | 765.1932 |
| rs7529220 | 1 | 22282619 | C | T | 0.0621 | 0.0098 | 1.98E-10 | -0.0051 | 0.0115 | 0.6555 | 1031.915 |
| rs6665642 | 1 | 154802139 | T | C | -0.062 | 0.0112 | 3.06E-08 | -0.0167 | 0.013 | 0.2 | 823.0411 |
| rs74832855 | 1 | 170171598 | G | A | 0.1216 | 0.018 | 1.43E-11 | 0.0052 | 0.0212 | 0.8068 | 1084.527 |
| rs72926475 | 2 | 86594487 | A | G | -0.0683 | 0.0102 | 2.37E-11 | -0.0151 | 0.012 | 0.2074 | 1037.036 |
| rs17060733 | 8 | 21866933 | C | T | -0.061 | 0.0103 | 2.64E-09 | -0.0236 | 0.0121 | 0.0511199 | 815.4107 |
| rs62521286 | 8 | 124551975 | G | A | 0.1202 | 0.0135 | 4.50E-19 | 0.0227 | 0.0159 | 0.1518 | 1847.252 |
| rs60212594 | 10 | 75414344 | C | G | -0.1176 | 0.0096 | 9.20E-35 | -0.0642 | 0.0113 | 1.19E-08 | 3522.484 |
| rs55734480 | 7 | 14372009 | A | G | 0.0548 | 0.0078 | 2.20E-12 | 0.0098 | 0.0092 | 0.2877 | 1160.308 |
| rs2860482 | 12 | 57105938 | C | A | -0.054 | 0.0076 | 1.21E-12 | -0.0256 | 0.009 | 4.46E-03 | 1197.285 |
| rs2359171 | 16 | 73053022 | A | T | 0.1746 | 0.0086 | 4.65E-91 | 0.0259 | 0.0102 | 0.01119 | 9196.112 |
| rs10773657 | 12 | 123327900 | A | C | -0.0575 | 0.0103 | 2.54E-08 | -0.0283 | 0.0123 | 0.0209802 | 811.4888 |
| rs6560886 | 12 | 133150210 | C | T | 0.051 | 0.009 | 1.49E-08 | -0.0152 | 0.0104 | 0.1443 | 895.3615 |
| rs9506925 | 13 | 23368943 | T | C | 0.0449 | 0.0075 | 2.72E-09 | 0.0041 | 0.0089 | 0.643899 | 813.8905 |
| rs2738413 | 14 | 64679960 | G | A | -0.0778 | 0.0067 | 2.55E-31 | -0.0034 | 0.0078 | 0.6624 | 3128.905 |
| rs74022964 | 15 | 73677264 | T | C | 0.1132 | 0.009 | 3.51E-36 | -0.0019 | 0.0108 | 0.8563 | 3508.443 |
| rs464901 | 22 | 18597502 | C | T | -0.0508 | 0.0072 | 1.53E-12 | -0.0026 | 0.0085 | 0.758 | 1187.149 |
| rs12426679 | 12 | 76237987 | T | C | -0.0391 | 0.0067 | 4.95E-09 | -8.00E-04 | 0.0079 | 0.92 | 786.1378 |
| rs4963776 | 12 | 24779491 | T | G | -0.0913 | 0.0088 | 1.84E-25 | -0.0214 | 0.0102 | 0.0364502 | 2565.962 |
| rs12245149 | 10 | 65321147 | A | C | -0.047 | 0.0067 | 1.66E-12 | -0.0143 | 0.0079 | 0.0693506 | 1136.706 |
| rs80056983 | 10 | 105509902 | T | C | 0.1287 | 0.0101 | 1.68E-37 | 0.0113 | 0.0121 | 0.3542 | 3516.334 |
| rs6994744 | 8 | 141740868 | C | A | 0.0405 | 0.0066 | 1.09E-09 | 0.011 | 0.0078 | 0.1597 | 846.035 |
| rs883079 | 12 | 114793240 | T | C | 0.0981 | 0.0074 | 2.84E-40 | 0.0176 | 0.0087 | 0.0439097 | 4123.156 |
| rs10141892 | 14 | 35184323 | C | T | -0.0452 | 0.0068 | 2.95E-11 | 0.0215 | 0.008 | 6.98E-03 | 1024.808 |
| rs55985730 | 7 | 128417044 | G | T | 0.0867 | 0.0149 | 5.24E-09 | 0.0094 | 0.0169 | 0.579499 | 874.7912 |
| rs337705 | 5 | 113737062 | G | T | 0.0564 | 0.0068 | 1.63E-16 | 0.005 | 0.0082 | 0.5424 | 1539.182 |
| rs34969716 | 6 | 18210109 | A | G | 0.0702 | 0.0078 | 1.60E-19 | 0.0012 | 0.0092 | 0.8961 | 2158.569 |
| rs34080181 | 3 | 66454191 | A | G | -0.0446 | 0.0069 | 1.28E-10 | -0.0028 | 0.0083 | 0.734301 | 966.1089 |
| rs12188351 | 5 | 168386089 | A | G | 0.0865 | 0.0145 | 2.52E-09 | 0.0222 | 0.0171 | 0.1923 | 821.6067 |
| rs1563304 | 17 | 44874453 | T | C | 0.0644 | 0.0092 | 2.56E-12 | 0.0277 | 0.0108 | 0.0102601 | 1252.592 |
| rs9953366 | 18 | 46474192 | C | T | 0.049 | 0.0073 | 1.82E-11 | 0.0259 | 0.0086 | 2.50E-03 | 1107.024 |
| rs140185678 | 16 | 2003016 | A | G | 0.1659 | 0.0218 | 2.43E-14 | 0.0759 | 0.0245 | 1.95E-03 | 1925.357 |
| rs77316573 | 16 | 2265271 | T | C | 0.0529 | 0.0089 | 3.26E-09 | -0.0039 | 0.0104 | 0.708 | 920.8044 |
| rs7225165 | 17 | 1309850 | A | G | -0.0655 | 0.0111 | 3.20E-09 | -0.0342 | 0.013 | 8.34E-03 | 889.37 |
| rs6838973 | 4 | 111765495 | T | C | -0.1514 | 0.0067 | 1.03E-111 | -0.0284 | 0.008 | 3.67E-04 | 11780.74 |
| rs6747542 | 2 | 70106832 | C | T | -0.0554 | 0.0067 | 1.10E-16 | -0.0278 | 0.0079 | 3.94E-04 | 1576.194 |
| rs4073778 | 1 | 116297758 | A | C | 0.0486 | 0.0067 | 4.96E-13 | -0.006 | 0.008 | 0.4558 | 1198.904 |
| rs11264280 | 1 | 154862952 | T | C | 0.1347 | 0.0071 | 3.07E-79 | 0.0252 | 0.0084 | 2.64E-03 | 8376.037 |
| rs3820888 | 2 | 201180023 | C | T | 0.0684 | 0.0068 | 5.75E-24 | 0.0366 | 0.008 | 4.71E-06 | 2304.25 |
| rs10804493 | 3 | 111554426 | A | G | 0.0558 | 0.007 | 1.63E-15 | 0.0126 | 0.0082 | 0.1242 | 1461.494 |
| rs6771054 | 3 | 89489529 | C | T | -0.0457 | 0.0068 | 2.42E-11 | -0.0014 | 0.0081 | 0.8584 | 1037.39 |
| rs1838747 | 5 | 114426668 | G | A | 0.0391 | 0.0067 | 4.13E-09 | 0.0149 | 0.0078 | 0.0571202 | 788.5107 |
| rs10213171 | 4 | 148937537 | G | C | 0.091 | 0.0134 | 1.32E-11 | 0.0362 | 0.0159 | 0.0230701 | 977.3322 |
| rs1278493 | 3 | 135814009 | A | G | -0.0389 | 0.0068 | 8.77E-09 | -0.0146 | 0.0079 | 0.0638102 | 767.5263 |
| rs9997349 | 4 | 111473366 | A | G | -0.0668 | 0.0086 | 5.40E-15 | -0.0231 | 0.0102 | 0.0244901 | 1494.406 |
| rs73041705 | 3 | 24463235 | C | T | -0.0443 | 0.0073 | 1.55E-09 | -0.0148 | 0.0086 | 0.0860498 | 847.9208 |
| rs11102343 | 1 | 112382671 | A | G | 0.0663 | 0.0111 | 2.23E-09 | 0.0191 | 0.0131 | 0.1452 | 791.4734 |
| rs56201652 | 7 | 92278116 | A | G | -0.0531 | 0.0075 | 1.74E-12 | -0.0257 | 0.0089 | 3.73E-03 | 1138.945 |
| rs6462079 | 7 | 28415827 | A | G | 0.0466 | 0.0076 | 8.79E-10 | 0.0025 | 0.0091 | 0.785301 | 901.7798 |
| rs3951016 | 6 | 118559658 | A | T | 0.0648 | 0.0067 | 2.15E-22 | -0.0144 | 0.0079 | 0.0671104 | 2154.197 |
| rs13195459 | 6 | 122403559 | A | G | -0.0623 | 0.007 | 4.15E-19 | -0.0043 | 0.0082 | 0.6023 | 1850.303 |
| rs72700114 | 1 | 170193825 | C | G | 0.2021 | 0.013 | 3.29E-54 | 0.0591 | 0.0152 | 9.48E-05 | 5918.607 |
| rs76097649 | 11 | 128764570 | A | G | 0.1151 | 0.0124 | 1.26E-20 | 0.0083 | 0.0145 | 0.566301 | 2315.737 |
| rs71454237 | 12 | 70013415 | A | G | -0.062 | 0.0084 | 1.78E-13 | -0.0119 | 0.0099 | 0.2269 | 1311.829 |
| rs7578393 | 2 | 26165528 | T | C | 0.0614 | 0.0088 | 2.42E-12 | -0.0035 | 0.0102 | 0.7323 | 1263.662 |
| rs10458660 | 10 | 77936576 | G | A | 0.0537 | 0.0087 | 6.78E-10 | 0.0121 | 0.0104 | 0.2448 | 849.731 |
| rs10821415 | 9 | 97713459 | A | C | 0.0821 | 0.0067 | 2.92E-34 | 0.0294 | 0.0079 | 2.18E-04 | 3380.472 |
| rs4935786 | 11 | 121661507 | A | T | -0.0463 | 0.0079 | 4.85E-09 | -0.0191 | 0.0094 | 0.0413 | 866.3047 |
| rs28631169 | 14 | 23888183 | T | C | 0.0522 | 0.0084 | 5.35E-10 | -0.0016 | 0.01 | 0.8752 | 893.523 |
| rs2759301 | 15 | 80994288 | A | G | 0.039 | 0.0067 | 5.04E-09 | 0.0287 | 0.0092 | 1.78E-03 | 777.9582 |
| rs12604076 | 17 | 76773638 | C | T | -0.0365 | 0.0066 | 3.63E-08 | -0.0306 | 0.0078 | 9.25E-05 | 685.7543 |
| rs2274115 | 9 | 139094773 | G | A | 0.0487 | 0.0076 | 1.69E-10 | 0.0202 | 0.009 | 0.0254203 | 1027.259 |
| rs35963991 | 8 | 11495702 | T | G | 0.0525 | 0.0095 | 2.80E-08 | 0.0288 | 0.0112 | 0.0100099 | 722.6328 |
| rs7789146 | 7 | 150661409 | A | G | -0.0584 | 0.0087 | 2.12E-11 | -0.0322 | 0.0103 | 1.78E-03 | 1033.013 |
| rs79187193 | 1 | 147255831 | A | G | -0.1162 | 0.0153 | 3.15E-14 | -0.0178 | 0.0172 | 0.301 | 1495.997 |
| rs6689306 | 1 | 154395946 | G | A | -0.046 | 0.0068 | 1.36E-11 | -0.0234 | 0.008 | 3.51E-03 | 1058.537 |
| rs577676 | 1 | 170587340 | T | C | -0.0923 | 0.0067 | 1.62E-43 | -0.0146 | 0.0079 | 0.0655194 | 4342.338 |
| rs2885697 | 1 | 41544279 | T | G | -0.0439 | 0.007 | 2.88E-10 | -0.0129 | 0.0082 | 0.1165 | 906.848 |
| rs6596717 | 5 | 106427609 | A | C | -0.0404 | 0.0068 | 3.00E-09 | 0.0037 | 0.008 | 0.6446 | 804.8427 |
| rs2040862 | 5 | 137419989 | T | C | 0.1084 | 0.0087 | 1.08E-35 | 0.0269 | 0.0103 | 8.81E-03 | 3548.985 |
| rs2031522 | 6 | 87821501 | G | A | -0.0436 | 0.0068 | 1.47E-10 | -0.0325 | 0.0081 | 6.04E-05 | 920.7361 |
| rs6790396 | 3 | 38771925 | G | C | 0.0627 | 0.0068 | 2.40E-20 | 0.0116 | 0.0079 | 0.1433 | 1955.416 |
| rs7650482 | 3 | 12841804 | G | A | 0.0711 | 0.007 | 1.79E-24 | 0.022 | 0.0082 | 7.40E-03 | 2406.58 |
| rs2288327 | 2 | 179411665 | G | A | 0.0919 | 0.0089 | 7.26E-25 | 0.0019 | 0.0107 | 0.8596 | 2302.46 |
| rs28387148 | 2 | 127433465 | T | C | 0.0741 | 0.0113 | 6.25E-11 | 0.029 | 0.0131 | 0.02618 | 1065.813 |
| rs7508 | 8 | 17913970 | A | G | 0.0711 | 0.0075 | 1.69E-21 | 0.012 | 0.0088 | 0.171 | 2146.436 |
| rs11773845 | 7 | 116191301 | A | C | 0.1054 | 0.0067 | 2.39E-55 | 0.0227 | 0.0079 | 4.19E-03 | 5588.159 |
| rs11191116 | 10 | 103555611 | T | C | -0.041 | 0.007 | 4.42E-09 | 0.0119 | 0.0083 | 0.1516 | 786.9456 |
| rs775498 | 12 | 70071513 | G | A | 0.0423 | 0.0074 | 1.05E-08 | 0.003 | 0.0087 | 0.7304 | 743.8983 |
| rs74884082 | 14 | 73249419 | T | C | -0.0493 | 0.0078 | 3.48E-10 | -0.0153 | 0.0092 | 0.0977305 | 939.1377 |
| rs35569628 | 13 | 113872712 | C | T | -0.0452 | 0.008 | 1.38E-08 | -0.0242 | 0.0095 | 0.01048 | 730.3467 |
| rs11156751 | 14 | 32990437 | C | T | 0.0719 | 0.0077 | 6.94E-21 | 0.029 | 0.009 | 1.27E-03 | 2177.803 |
| rs3176326 | 6 | 36647289 | A | G | -0.0626 | 0.0085 | 1.42E-13 | -0.0577 | 0.0102 | 1.33E-08 | 1285.519 |
| rs74500426 | 4 | 174642789 | T | G | -0.0921 | 0.0127 | 4.29E-13 | -0.0091 | 0.0144 | 0.527 | 1235.479 |

**Supplementary File-Table 3.**
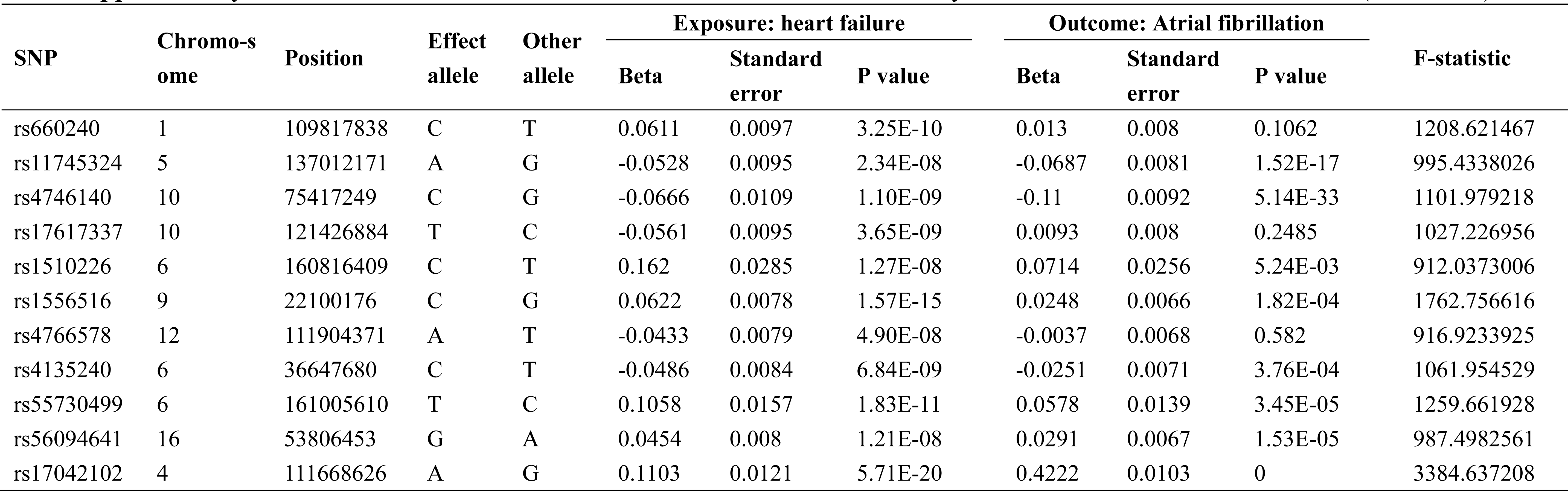
SNPs used to construct the instruments for MR analysis heart failure and atrial fibrillation (nSNPs=11).

**Supplementary File-Table 4.** Sensitivity analysis of association between atrial fibrillation.

| Outcomes | Exposures | nSNPs | beta | se | P value | Cochrane Q<br>test pvalue | MR-Egger<br>intercept<br>pvalue | Global<br>test<br>pvalue | Distortion<br>test pvalue |
| --- | --- | --- | --- | --- | --- | --- | --- | --- | --- |
| Heart failure | Atrial fibrillation | 99 | 0.2153007 | 0.01924315 | 4.65E-29 | 5.93E-10 | 0.090 | <0.001 | 0.675 |
| Atrial fibrillation | Heart failure | 9 | 1.3547512 | 0.50191937 | 6.95E-03 | 1.28E-215 | 0.171 | <0.001 | 1 |
| Heart failure | Waist circumference | 59 | 0.02550309 | 0.05473641 | 0.6412688 | 0.008 | 0.516 | 0.008 | NA |
|  | Body mass index | 448 | 0.5518767 | 0.03156728 | 1.95E-68 | 1.48E-09 | 0.153 | <0.001 | 0.874 |
|  | Systolic blood pressure | 398 | 0.02107779 | 0.002081169 | 4.16E-24 | 8.78E-23 | 0.391 | <0.001 | 0.437 |
|  | Diastolic blood pressure | 391 | 0.02960439 | 0.003640814 | 4.25E-16 | 7.12E-27 | 0.484 | <0.001 | 0.815 |
|  | HbA1C | 188 | -0.06584305 | 0.06512364 | 0.311994 | 1.41E-06 | 0.310 | <0.001 | 0.829 |
|  | Fasting glucose | 19 | 0.1582335 | 0.06474746 | 0.01453122 | 0.718 | 0.618 | 0.745 | - |
|  | Fasting insulin | 4 | 0.19998547 | 0.3942165 | 0.6119453 | 0.016 | 0.723 | 0.062 | - |
|  | Smoking | 73 | 0.2372768 | 0.0484872 | 9.90E-07 | 0.005 | 0.863 | 0.001 | 0.754 |
|  | Alcoholic drinking | 30 | 0.133789 | 0.1255667 | 0.28665816 | 3.03E-04 | 0.971 | <0.001 | 0.521 |
|  | Physical activity | 11 | 0.0529421 | 0.1304013 | 0.6847473 | 0.028 | 0.901 | 0.029 | NA |
|  | PM2.5 exposure | 6 | 1.26083038 | 0.8335022 | 0.1303584 | 1.05E-09 | 0.536 | <0.001 | <0.001 |
|  | Coronary heart disease | 38 | 0.2909573 | 0.02736351 | 2.09E-26 | 3.03E-07 | 0.336 | <0.001 | 0.767 |
|  | Myocardial infarction | 23 | 0.300232 | 0.03399264 | 1.03E-18 | 1.67E-06 | 0.339 | <0.001 | 0.746 |
| Atrial fibrillation | Waist circumference | 63 | 0.3196562 | 0.07827214 | 4.43E-05 | 3.76E-30 | 0.301 | <0.001 | 0.689 |
|  | Body mass index | 491 | 0.3453513 | 0.03091049 | 5.55E-29 | 7.80E-41 | 0.892 | <0.001 | 0.425 |
|  | Systolic blood pressure | 438 | 0.01608193 | 0.00214104 | 5.86E-14 | 1.67E-92 | 0.919 | <0.001 | 0.838 |
|  | Diastolic blood pressure | 442 | 0.02904597 | 0.003639805 | 1.46E-15 | 7.19E-98 | 0.188 | <0.001 | 0.691 |
|  | HbA1C | 225 | 0.4113814 | 0.07106406 | 7.09E-09 | 6.95E-49 | 0.078 | <0.001 | 0.933 |
|  | Fasting glucose | 22 | -0.034893326 | 0.07319565 | 0.6335664 | 1.79E-04 | 0.212 | 0.001 | 0.442 |
|  | Fasting insulin | 3 | -0.21556932 | 0.2216626 | 0.3307964 | 0.316 | 0.419 | NA | NA |
|  | Smoking | 85 | 0.08767079 | 0.04059599 | 0.03080424 | 0.005 | 0.863 | <0.001 | 0.639 |
|  | Alcoholic drinking | 33 | 0.1280652 | 0.09928672 | 0.19710177 | 0.001 | 0.906 | 0.001 | 0.582 |
|  | Physical activity | 11 | 0.007033304 | 0.125875 | 0.9554412 | 0.028 | 0.901 | 0.003 | 0.709 |
|  | PM2.5 exposure | 7 | 0.3981797 | 0.3918176 | 0.3095163 | 1.05E-09 | 0.536 | 0.003 | 0.943 |
|  | Coronary heart disease | 39 | 0.10563334 | 0.02680572 | 8.12E-05 | 3.03E-07 | 0.336 | <0.001 | 0.826 |
|  | Myocardial infarction | 24 | 0.1030076 | 0.02417212 | 2.03E-05 | 0.001 | 0.039 | 0.004 | 0.652 |

**Supplementary File-Fig 1.**
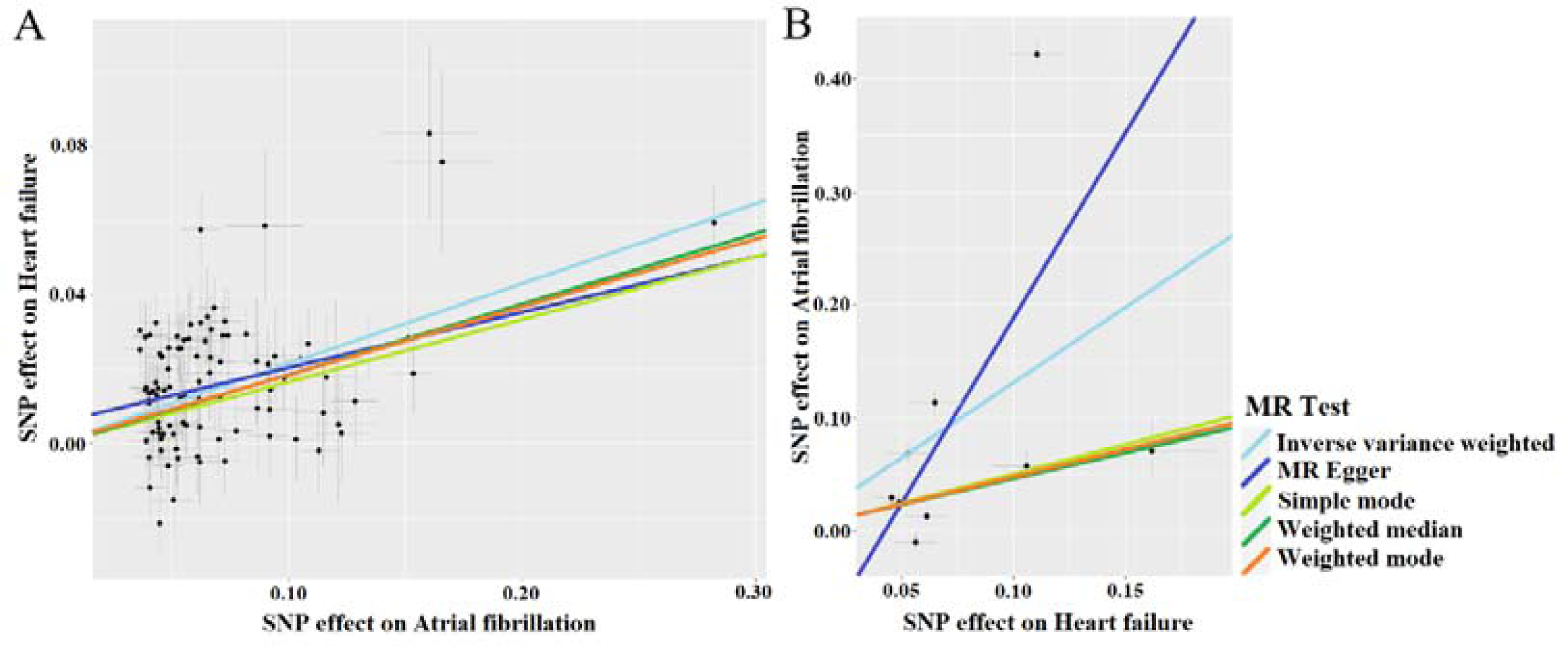
Individual estimates of a bidirectional MR analysis of atrial fibrillation and heart failure. (A: Genetic variation of atrial fibrillation in individual estimates of heart failure; B: Genetic variation of heart failure to individual estimates of atrial fibrillation)

**Supplementary File-Fig 2.**
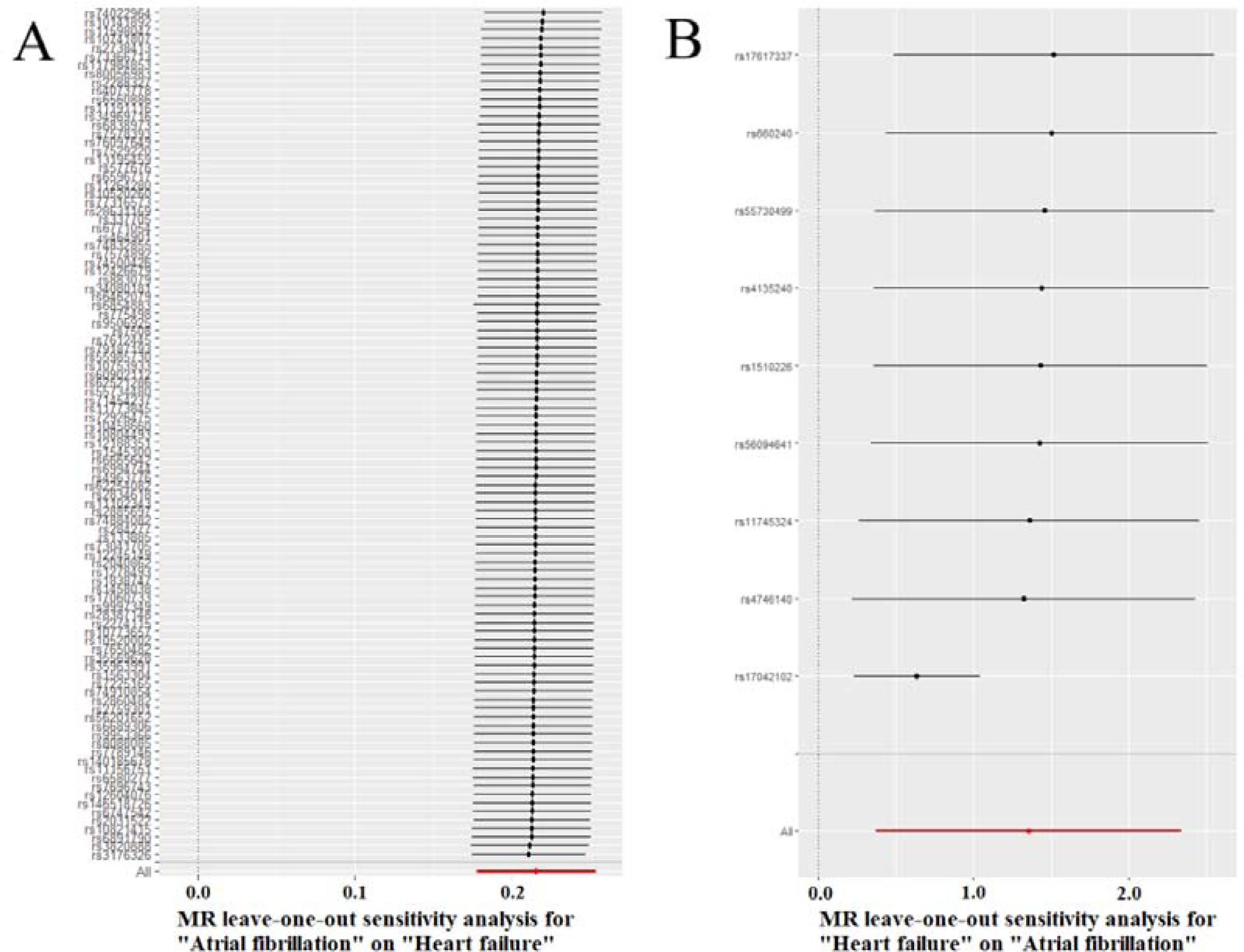
Leave-one-out method analysis results of bidirectional MR association between atrial fibrillation and heart failure. (A: Atrial fibrillation → Heart failure; B: Heart failure → Atrial fibrillation)

## Notes

### Competing Interest Statement

The authors have declared no competing interest.

### Funding Statement

This work was supported by the Natural Science Foundation of Fujian Province (grant no. 2022J01706) and the Start-up Fund for high-level talents of Fujian Medical University (XRCZX2021026) to Dr. Wuqing Huang. The funder had no role in study design, data collection and interpretation, or the decision to submit the work for publication.

### Author Declarations

All these studies had been approved by the relevant institutional review boards and informed consent had been obtained for all participants per the original study protocols.

